# Parental Preferences and Reasons for COVID-19 Vaccination Among Their Children

**DOI:** 10.1101/2022.10.20.22281313

**Authors:** Neil K. R. Sehgal, Benjamin Rader, Autumn Gertz, Christina M. Astley, John S. Brownstein

## Abstract

**Background:** COVID-19 vaccination rates among children have stalled, while new coronavirus strains continue to emerge. To improve child vaccination rates, policymakers must better understand parental preferences and reasons for COVID-19 vaccination among their children.

**Methods and Findings:** Cross-sectional surveys were administered online to 30,174 US parents with at least one child of COVID-19 vaccine eligible age (5-17 years) between January 1 and May 9, 2022. Participants self-reported willingness to vaccinate their child and reasons for hesitancy, and answered additional questions about demographics, pandemic related behavior, and vaccination status. Willingness to vaccinate a child for COVID-19 was strongly associated with parental vaccination status (multivariate odds ratio 97.9, 95% confidence interval 86.9-111.0). The majority of fully vaccinated (86%) and unvaccinated (84%) parents reported concordant vaccination preferences for their eligible child. Age and education had differing relationships by vaccination status, with higher age and education positively associated with willingness among vaccinated parents. Among all parents hesitant to vaccinate their children, the two most frequently reported reasons were possible side effects (47%) and that vaccines are too new (44%). Among hesitant parents, parental vaccination status was inversely associated with reported lack of trust in government (p<.001) and scientists (p<.001). Cluster analysis identified three groups of hesitant parents based on their reasons for hesitance to vaccinate, with distinct concerns that may be obscured when analyzed in aggregate.

**Conclusion:** Factors associated with willingness to vaccinate children and reasons for hesitancy may inform targeted approaches to increase vaccination.

## Introduction

In the United States (US), children as young as 5 years old became eligible for the Pfizer-BioNTech COVID-19 vaccine in October 2021 [1]. Studies of the vaccine in real world settings have found it to be highly effective in preventing severe disease and reducing hospitalization among children and adolescents [2,3]. However, despite wide availability of vaccines in the US, many children remain unvaccinated and vaccine uptake appears to have slowed [4]. On May 25, 2022, just 51% of children ages 5-17 had received at least one dose of a COVID-19 vaccine. By September 21, 2022, rates had risen by just one percentage point to 52% [5]. This is a lower rate compared to coverage levels of many other similarly recommended immunizations seen in the past. For instance, in the 2018-2019 school year, 63% of children between 6 months and 18 years received a flu vaccine [6]. Likewise, the coverage rate for the meningococcal conjugate (MenACWY), a disease with similarly rare but severe effects, was 89% among adolescents in 2020 [7].

According to the Centers for Disease Control and Prevention (CDC), as of September 2022, there have been over 15 million COVID-19 cases, over 12,200 laboratory-confirmed COVID-19 associated hospitalizations, and over 1,700 COVID-19 deaths among children in the United States (US) [8,9]. While adults have faced much higher acute morbidity and mortality, long term impacts of the disease on children are still unclear. Recent estimates from the United Kingdom suggest that around 5% of all secondary school students have experienced Post-COVID Conditions [10]. Additionally, children have faced far-reaching secondary effects from the pandemic, including disrupted healthcare services, routine immunizations, and education, and an increased prevalence of mental health disorders such as anxiety and depression [11].

Given the large disparities in vaccine coverage between COVID-19 and routine pediatric immunizations, increasing COVID-19 vaccination rates among children may require new strategies. Much prior work has been devoted to understanding the COVID-19 hesitancy among parents. Prior studies have found vaccinated parents much more likely to vaccinate their children and that side effects and vaccine safety are two of the most common reasons for concern [4, 12-22]. However, prior studies often analyzed all parents as a single cohort, rather than investigating this heterogeneity. Additionally, prior research has been limited by potentially non-representative samples. In this large, diverse, and nationally representative study, we aim to identify the demographic factors associated with parental preferences for COVID-19 vaccination among their children and reasons for hesitancy. We also examine heterogeneity among vaccinated and unvaccinated parents through the inclusion of numerous relevant predictor variables and detailed information on reasons for hesitancy.

## Methods

To understand hesitancy for child vaccination against COVID-19, we administered an online survey focusing on parents with children ages 5-17, and analyzed responses using survey weights, logistic regression, and cluster analysis.

### Participants

Each day, over 2 million individuals complete online surveys using the SurveyMonkey platform by Momentive [23]. Between January 1 and May 9, 2022, a random subset of US SurveyMonkey participants were invited to complete an additional questionnaire on COVID-19. There were no financial incentives for completing the survey, with 217,023 of those invited choosing to participate. Primary analysis restricted to parents ages 18+ who self-reported eligible children ages 5 to 17 (i.e. excluded parents with any ineligible children <5 yr). To evaluate the impact of excluding parents of children < 5 years, which could be a proxy for family size, we conducted a sensitivity analysis without this exclusion (Appendix Table 1).

### Survey Questions

Questions were selected from a previously validated web-survey [24,25]. Participants answered questions about demographics, pandemic related behavior, and vaccination status. Participants who received at least one dose of the vaccine were classified as vaccinated. Participants were asked if they had children of the following ages: under 5, 5 to 11, 12 to 15, and 16 to 17. For each age group that parents had children of, they were additionally asked if they were willing to have their child(ren) of that age group vaccinated against COVID-19. Participants could respond with “yes”, “no”, or “not sure”. To understand parents with clear preferences we restricted our analysis to parents always willing or unwilling to vaccinate all of their children. Participants who were unwilling to vaccinate at least one of their children were asked to select up to 20 non-exclusive l reasons for their hesitancy from a list (Appendix Table 2).

### Analyses

Using the latest estimates from the U.S. Census Bureau’s American Community Survey, survey weights were applied to reflect the US adult population in demographic composition and political beliefs [26]. Census derived weights accounted for age, race, sex, education, and geography. To account for political beliefs, an additional smoothing parameter for political party identification was included based on aggregates of SurveyMonkey research surveys [24].

Weighted multivariate logistic regression models were used to assess the association between parental characteristics and willingness to vaccinate their children. The estimation equation is:

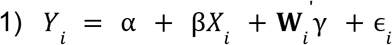

where *Y*_i_ is a binary variable for the willingness of parent i to vaccinate their child(ren), *X* _i_ is a dummy variable equal to 1 if the parent i is vaccinated, and **W** _i_ is a vector of parent level factors.

To assess heterogeneity in correlates of willingness between vaccinated and unvaccinated parents, we fit regressions separately for fully vaccinated and unvaccinated parents, as well as for the overall sample. We used KModes cluster analysis, an extension of KMeans for categorical data, to group hesitant parents into clusters according to their reasons for reluctance [27]. All statistical analyses were conducted in R version 4.1.2.

## Results

Of the 217,023 survey respondents, 3,301 were under the age of 18 and excluded. An additional 159,752 users were excluded for not having children 17 or under and 10,098 users were excluded for having children under the age of 5. Next, 9271 respondents were excluded for missing data on variables of interest. Lastly, 3,410 parents were removed who were always unsure about vaccinating their children, and 1,051 were removed for only intending to vaccinate some of their children, leaving a total sample of 30,140. After exclusion, the mean survey weight of participants was .98, leading to a weighted sample of 29,583.

Of the 29,583 surveyed parents, 7,654 (26%) were unvaccinated, 2,697 (9%) were partially vaccinated, 9,713 (33%) were fully vaccinated, and 9,519 (32%) were fully vaccinated and boosted. A total of 19,586 (66%) parents expressed willingness to vaccinate their child. Parent characteristics are listed in Table 1. About half of participants were female, and a majority were White, religious, employed, and insured. A plurality of parents were ages 40-49 years, and had a household income under $49,999. Appendix Table 3 displays summary characteristics of parents who were always unsure or held inconsistent preferences across their children.

**Table 1.** Characteristics of Studied Participants

|  | Weighted (N=29,583) | Unweighted (N=30,140) |
| --- | --- | --- |
|  | n (%) | n (%) |
| Gender |  |  |
| Female | 15,304 (52%) | 18,915 (63%) |
| Male | 13,949 (47%) | 10,885 (36%) |
| Transgender or Nonbinary | 330 (1%) | 340 (1%) |
| Age |  |  |
| 18-29 years | 1,686 (6%) | 1,050 (3%) |
| 30-39 years | 8,018 (27%) | 6,616 (22%) |
| 40-49 years | 12,090 (41%) | 13,206 (44%) |
| 50-64 years | 6,872 (23%) | 8,473 (28%) |
| 65+ years | 917 (3%) | 795 (3%) |
| Household Income |  |  |
| Under \$49,999 | 11,054 (37%) | 9,137 (30%) |
| \$50,000-\$99,999 | 8,384 (28%) | 8,237 (27%) |
| Over \$100,000 | 10,146 (34%) | 12,766 (42%) |
| Race/Ethnicity |  |  |
| White, not Hispanic | 17,150 (58%) | 17,609 (58%) |
| Hispanic | 5,947 (20%) | 4,859 (16%) |
| Black | 5,947 (20%) | 4,859 (16%) |
| Asian | 1,677 (6%) | 1,621 (5%) |
| Other | 1,169 (4%) | 1,346 (4%) |
| Education |  |  |
| High School or Less | 11,294 (38%) | 5,762 (19%) |
| Some College | 9,075 (31%) | 8,894 (30%) |
| College Graduate | 9,215 (31%) | 15,484 (51%) |
| Employment Status |  |  |
| Employed | 24,388 (82%) | 25,449 (84%) |
| Unemployed | 5,195 (18%) | 4,691 (16%) |
| Health Insurance |  |  |
| Insured | 27,253 (92%) | 28,403 (94%) |
| Uninsured | 2,331 (8%) | 1,737 (6%) |
| Self Reported Health |  |  |
| Fair/Poor | 2,388 (8%) | 2,274 (8%) |
| Good | 7,588 (26%) | 7,598 (25%) |
| Very good | 11,194 (38%) | 11,771 (39%) |
| Excellent | 8,413 (28%) | 8,497 (28%) |
| Religious Status |  |  |
| Religious | 22,484 (76%) | 22,536 (75%) |
| Atheist/Agnostic | 7,099 (24%) | 7,604 (25%) |
| Parent COVID-19 Vaccination |  |  |
| Johnson | 1,970 (7%) | 1,856 (6%) |
| mRNA | 19,959 (67%) | 22,213 (74%) |
| Unvaccinated | 7,654 (26%) | 6,071 (20%) |
| Have Child Age 5 to 11 Years | 15,357 (52%) | 14,679 (49%) |
| Have Child Age 12 to 15 Years | 14,188 (48%) | 14,783 (49%) |
| Have Child Age 16 to 17 Years | 9,828 (33%) | 10,667 (35%) |
| Political Party Affiliation |  |  |
| Republican | 9,712 (33%) | 7,504 (25%) |
| Democrat | 9,538 (32%) | 11,402 (38%) |
| Independent | 10,334 (35%) | 11,234 (37%) |
| When Will the Pandemic End? |  |  |
| Already Over | 6,541 (22%) | 5,615 (19%) |
| Less than three months | 1,662 (6%) | 1,585 (5%) |
| Between three months and one year | 5,526 (19%) | 6,070 (20%) |
| More than one year | 15,854 (54%) | 16,870 (56%) |
| Flu Vaccine Since June 2021 | 13,935 (47%) | 15,924 (53%) |

We observed a high degree of concordance in preferences, with 86% of vaccinated parents willing to have their children vaccinated and 85% of unvaccinated parents unwilling to have their children vaccinated. In a pooled regression of all parents, willingness to vaccinate children was independently associated with a number of factors (Table 2). Major predictors of parents’ preferences for child vaccination included being fully vaccinated and boosted (odds ratio [OR]: 106.0, 95% confidence interval [CI]: 93.9-120.0), Democrat political affiliation (OR: 4.22, CI: 3.82-4.66), and Asian race (OR: 3.07, CI: 2.56-3.71) (Figure 1).

**Table 2.**
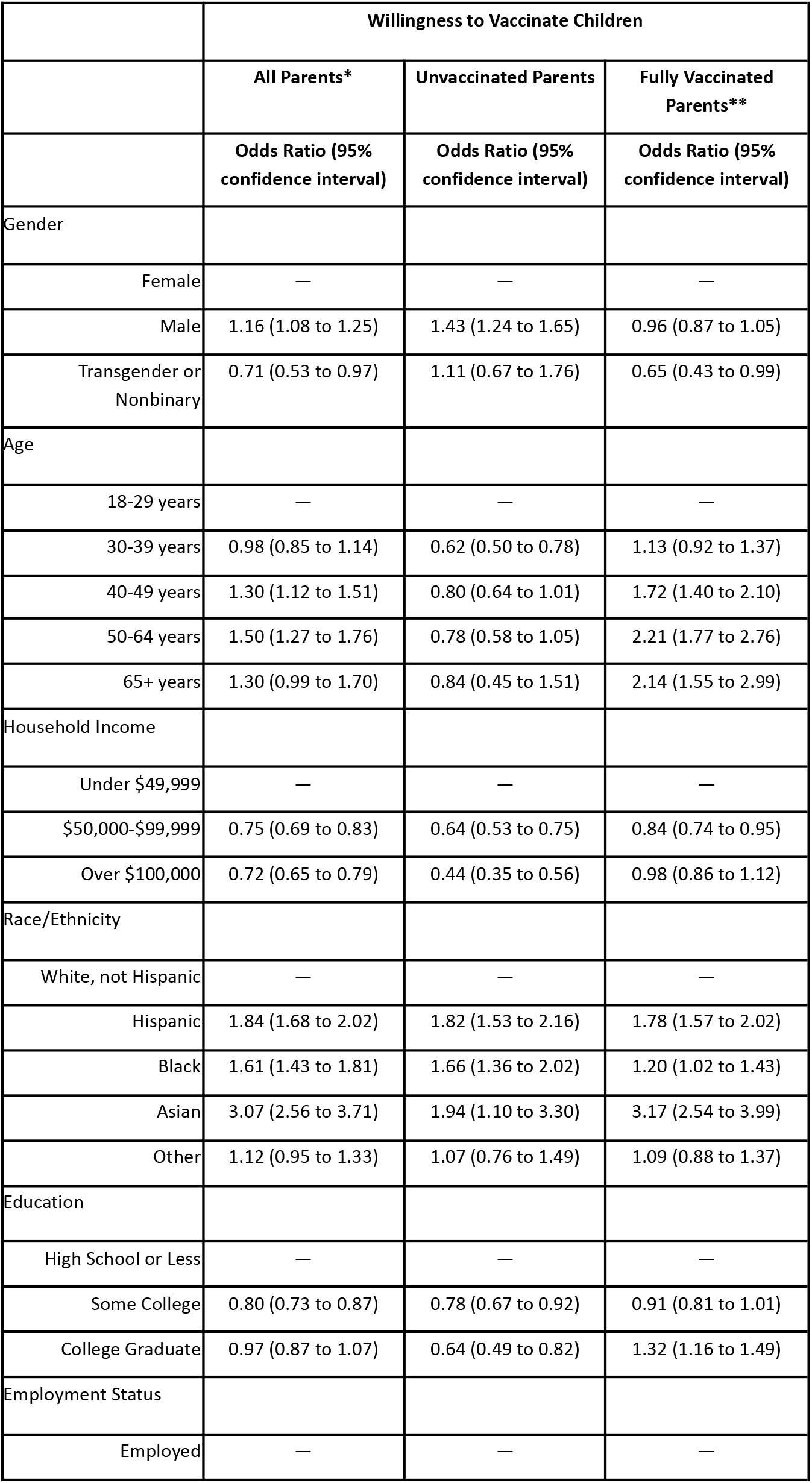
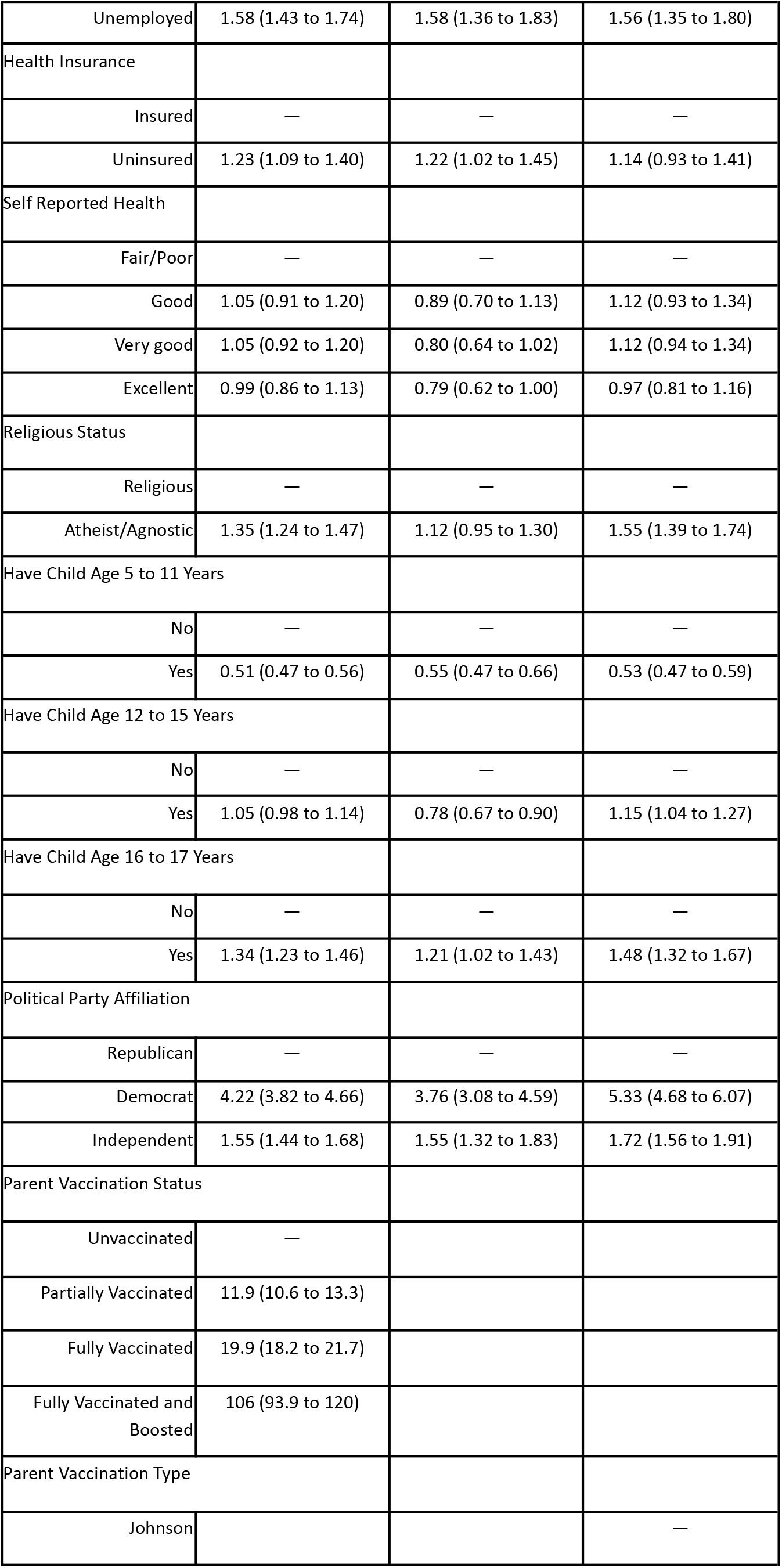
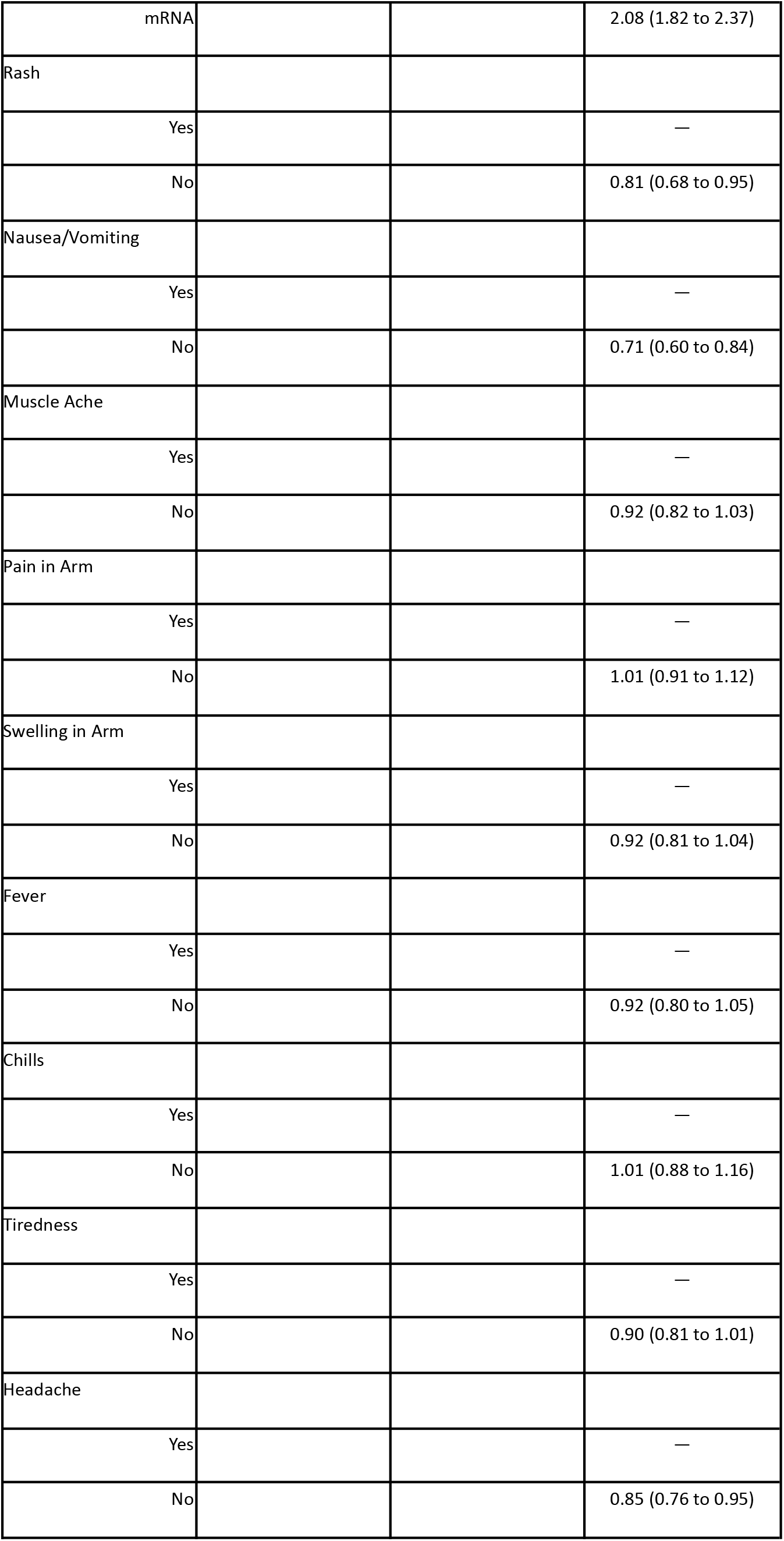
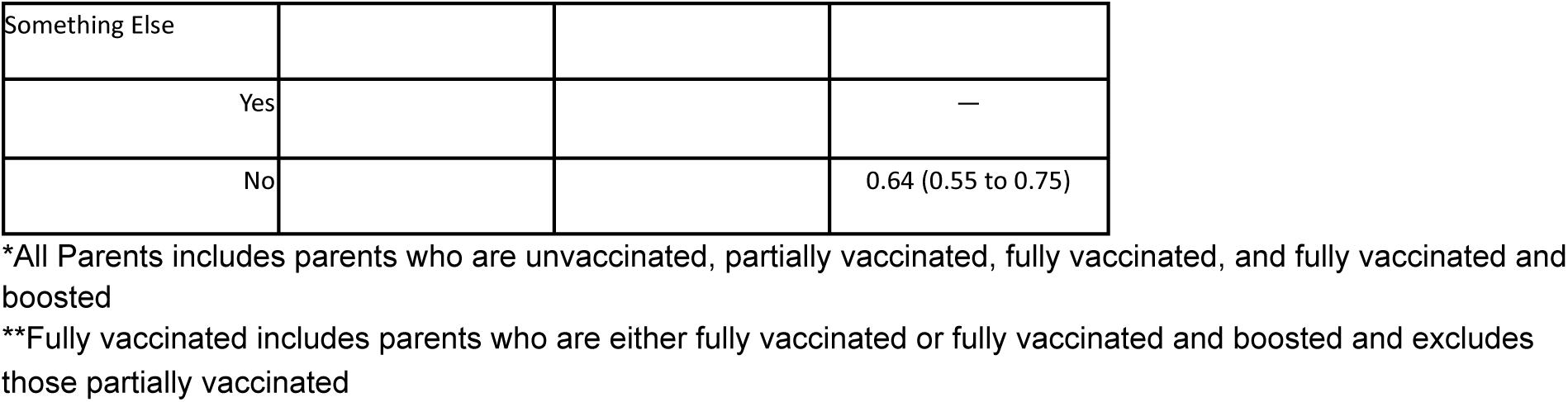
Multivariate Relationship between Parent Characteristics and Willingness to Vaccinate Children. *All Parents includes parents who are unvaccinated, partially vaccinated, fully vaccinated, and fully vaccinated and boosted **Fully vaccinated includes parents who are either fully vaccinated or fully vaccinated and boosted and excludes those partially vaccinated

|  | Willingness to Vaccinate Children |  |  |
| --- | --- | --- | --- |
|  | All Parents* | Unvaccinated Parents | Fully Vaccinated Parents** |
|  | Odds Ratio (95% confidence interval) | Odds Ratio (95% confidence interval) | Odds Ratio (95% confidence interval) |
| Gender |  |  |  |
| Female | — | — | — |
| Male | 1.16 (1.08 to 1.25) | 1.43 (1.24 to 1.65) | 0.96 (0.87 to 1.05) |
| Transgender or Nonbinary | 0.71 (0.53 to 0.97) | 1.11 (0.67 to 1.76) | 0.65 (0.43 to 0.99) |
| Age |  |  |  |
| 18-29 years | — | — | — |
| 30-39 years | 0.98 (0.85 to 1.14) | 0.62 (0.50 to 0.78) | 1.13 (0.92 to 1.37) |
| 40-49 years | 1.30 (1.12 to 1.51) | 0.80 (0.64 to 1.01) | 1.72 (1.40 to 2.10) |
| 50-64 years | 1.50 (1.27 to 1.76) | 0.78 (0.58 to 1.05) | 2.21 (1.77 to 2.76) |
| 65+ years | 1.30 (0.99 to 1.70) | 0.84 (0.45 to 1.51) | 2.14 (1.55 to 2.99) |
| Household Income |  |  |  |
| Under \$49,999 | — | — | — |
| \$50,000-\$99,999 | 0.75 (0.69 to 0.83) | 0.64 (0.53 to 0.75) | 0.84 (0.74 to 0.95) |
| Over \$100,000 | 0.72 (0.65 to 0.79) | 0.44 (0.35 to 0.56) | 0.98 (0.86 to 1.12) |
| Race/Ethnicity |  |  |  |
| White, not Hispanic | — | — | — |
| Hispanic | 1.84 (1.68 to 2.02) | 1.82 (1.53 to 2.16) | 1.78 (1.57 to 2.02) |
| Black | 1.61 (1.43 to 1.81) | 1.66 (1.36 to 2.02) | 1.20 (1.02 to 1.43) |
| Asian | 3.07 (2.56 to 3.71) | 1.94 (1.10 to 3.30) | 3.17 (2.54 to 3.99) |
| Other | 1.12 (0.95 to 1.33) | 1.07 (0.76 to 1.49) | 1.09 (0.88 to 1.37) |
| Education |  |  |  |
| High School or Less | — | — | — |
| Some College | 0.80 (0.73 to 0.87) | 0.78 (0.67 to 0.92) | 0.91 (0.81 to 1.01) |
| College Graduate | 0.97 (0.87 to 1.07) | 0.64 (0.49 to 0.82) | 1.32 (1.16 to 1.49) |
| Employment Status |  |  |  |
| Employed | — | — | — |
| Unemployed | 1.58 (1.43 to 1.74) | 1.58 (1.36 to 1.83) | 1.56 (1.35 to 1.80) |
| Health Insurance |  |  |  |
| Insured | — | — | — |
| Uninsured | 1.23 (1.09 to 1.40) | 1.22 (1.02 to 1.45) | 1.14 (0.93 to 1.41) |
| Self Reported Health |  |  |  |
| Fair/Poor | — | — | — |
| Good | 1.05 (0.91 to 1.20) | 0.89 (0.70 to 1.13) | 1.12 (0.93 to 1.34) |
| Very good | 1.05 (0.92 to 1.20) | 0.80 (0.64 to 1.02) | 1.12 (0.94 to 1.34) |
| Excellent | 0.99 (0.86 to 1.13) | 0.79 (0.62 to 1.00) | 0.97 (0.81 to 1.16) |
| Religious Status |  |  |  |
| Religious | — | — | — |
| Atheist/Agnostic | 1.35 (1.24 to 1.47) | 1.12 (0.95 to 1.30) | 1.55 (1.39 to 1.74) |
| Have Child Age 5 to 11 Years |  |  |  |
| No | — | — | — |
| Yes | 0.51 (0.47 to 0.56) | 0.55 (0.47 to 0.66) | 0.53 (0.47 to 0.59) |
| Have Child Age 12 to 15 Years |  |  |  |
| No | — | — | — |
| Yes | 1.05 (0.98 to 1.14) | 0.78 (0.67 to 0.90) | 1.15 (1.04 to 1.27) |
| Have Child Age 16 to 17 Years |  |  |  |
| No | — | — | — |
| Yes | 1.34 (1.23 to 1.46) | 1.21 (1.02 to 1.43) | 1.48 (1.32 to 1.67) |
| Political Party Affiliation |  |  |  |
| Republican | — | — | — |
| Democrat | 4.22 (3.82 to 4.66) | 3.76 (3.08 to 4.59) | 5.33 (4.68 to 6.07) |
| Independent | 1.55 (1.44 to 1.68) | 1.55 (1.32 to 1.83) | 1.72 (1.56 to 1.91) |
| Parent Vaccination Status |  |  |  |
| Unvaccinated | — |  |  |
| Partially Vaccinated | 11.9 (10.6 to 13.3) |  |  |
| Fully Vaccinated | 19.9 (18.2 to 21.7) |  |  |
| Fully Vaccinated and Boosted | 106 (93.9 to 120) |  |  |
| Parent Vaccination Type |  |  |  |
| Johnson |  |  | — |
| mRNA |  |  | 2.08 (1.82 to 2.37) |
| Rash |  |  |  |
| Yes |  |  | — |
| No |  |  | 0.81 (0.68 to 0.95) |
| Nausea/Vomiting |  |  |  |
| Yes |  |  | — |
| No |  |  | 0.71 (0.60 to 0.84) |
| Muscle Ache |  |  |  |
| Yes |  |  | — |
| No |  |  | 0.92 (0.82 to 1.03) |
| Pain in Arm |  |  |  |
| Yes |  |  | — |
| No |  |  | 1.01 (0.91 to 1.12) |
| Swelling in Arm |  |  |  |
| Yes |  |  | — |
| No |  |  | 0.92 (0.81 to 1.04) |
| Fever |  |  |  |
| Yes |  |  | — |
| No |  |  | 0.92 (0.80 to 1.05) |
| Chills |  |  |  |
| Yes |  |  | — |
| No |  |  | 1.01 (0.88 to 1.16) |
| Tiredness |  |  |  |
| Yes |  |  | — |
| No |  |  | 0.90 (0.81 to 1.01) |
| Headache |  |  |  |
| Yes |  |  | — |
| No |  |  | 0.85 (0.76 to 0.95) |
| Something Else |  |  |  |
| Yes |  |  | — |
| No |  |  | 0.64 (0.55 to 0.75) |

**Figure 1.**
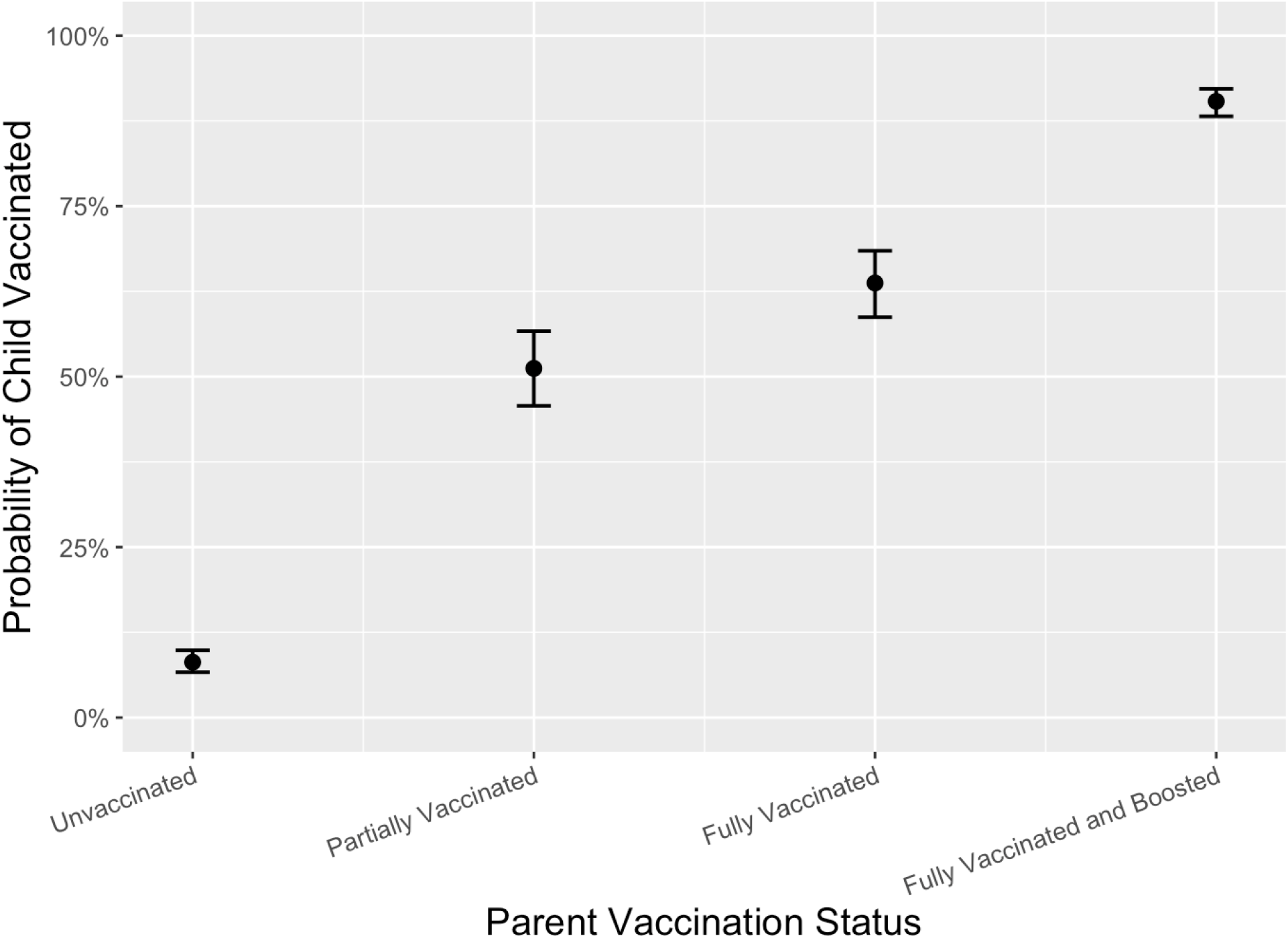
Predicted Probabilities of Vaccinating Child

Several correlates of willingness differed in our separated regression models of unvaccinated and fully vaccinated parents (Table 2). Among fully vaccinated parents, willingness was independently positively associated with older age, college education, atheism/agnosticism, receiving an mRNA vaccine, and no vaccine side effect of headache, nausea/vomiting, or rash. Among unvaccinated parents, willingness was independently positively associated with male gender, younger age, high school or less education, and a lack of health insurance.

The 9,998 parents who were unwilling to vaccinate their children listed several reasons for hesitancy (Appendix Table 2). Overall, parents were most concerned about potential side effects (47%) and the vaccine being too new (44%). Vaccinated and unvaccinated parents differed on several concerns. For example, unvaccinated parents were much more likely to list a lack of trust in government (41% to 21%, p<.001) and a lack of trust in scientists (34% to 19%, p<.001) as reasons for hesitancy. Parents with older children are more likely to say the decision to vaccinate should be the child’s decision when they are of appropriate age while parents with younger children are more likely to say their children are too young to be vaccinated (Appendix Table 4).

Cluster analysis of hesitant parents’ reasons indicated that 3 clusters achieved a good balance of interpretability and cluster quality. These clusters differ significantly on both reasons for hesitancy and parent characteristics (Table 3). For instance, Cluster 1 parents (32%) had a median of 8.0 reasons for hesitancy, were very likely to lack trust in government and scientists, were very likely to think the vaccine was too new and worry about potential side effects, and were likely to view the pandemic as already being over. Cluster 2 parents make up the majority of all hesitant parents (56%). Parents in this cluster had a median of 1.0 reasons for hesitancy and a majority (53%) had a high school or less education. Cluster 3 parents were the smallest group, had a median of 4.0 reasons for hesitancy, and were very likely to think the vaccine was too new and worry about potential side effects.

**Table 3.** Summary Characteristics by Clusters of Hesitant Parent

|  | *All Hesitant Parents | Cluster 1 |  | Cluster 2 |  | Cluster 3 |  |
| --- | --- | --- | --- | --- | --- | --- | --- |
|  | Overall, N = 9,998 | Unvaccinated, N = 2,342 | Vaccinated**, N = 847 | Unvaccinated, N = 3,400 | Vaccinated**, N = 2,176 | Unvaccinated, N = 733 | Vaccinated**, N = 500 |
| Characteristic |  |  |  |  |  |  |  |
| Gender |  |  |  |  |  |  |  |
| Female | 5,008/9,998 (50%) | 1,106/2,342 (47%) | 330/847 (39%) | 1,685/3,400 (50%) | 1,124/2,176 (52%) | 458/733 (62%) | 306/500 (61%) |
| Male | 4,830/9,998 (48%) | 1,192/2,342 (51%) | 497/847 (59%) | 1,662/3,400 (49%) | 1,030/2,176 (47%) | 262/733 (36%) | 187/500 (37%) |
| Transgender or Nonbinary | 159/9,998 (1.6%) | 44/2,342 (1.9%) | 20/847 (2.4%) | 53/3,400 (1.6%) | 22/2,176 (1.0%) | 13/733 (1.8%) | 7/500 (1.4%) |
| Age |  |  |  |  |  |  |  |
| 18-29 years | 859/9,998 (8.6%) | 119/2,342 (5.1%) | 35/847 (4.1%) | 394/3,400 (12%) | 233/2,176 (11%) | 54/733 (7.3%) | 25/500 (5.0%) |
| 30-39 years | 3,732/9,998 (37%) | 847/2,342 (36%) | 234/847 (28%) | 1,484/3,400 (44%) | 717/2,176 (33%) | 263/733 (36%) | 187/500 (38%) |
| 40-49 years | 3,757/9,998 (38%) | 930/2,342 (40%) | 367/847 (43%) | 1,116/3,400 (33%) | 844/2,176 (39%) | 306/733 (42%) | 194/500 (39%) |
| 50-64 years | 1,492/9,998 (15%) | 408/2,342 (17%) | 186/847 (22%) | 371/3,400 (11%) | 330/2,176 (15%) | 107/733 (15%) | 89/500 (18%) |
| 65+ years | 158/9,998 (1.6%) | 39/2,342 (1.7%) | 25/847 (3.0%) | 35/3,400 (1.0%) | 52/2,176 (2.4%) | 4/733 (0.5%) | 4/500 (0.7%) |
| Household Income |  |  |  |  |  |  |  |
| Under \$49,999 | 4,135/9,998 (41%) | 798/2,342 (34%) | 182/847 (22%) | 1,865/3,400 (55%) | 862/2,176 (40%) | 273/733 (37%) | 153/500 (31%) |
| \$50,00-\$99,999 | 3,026/9,998 (30%) | 733/2,342 (31%) | 262/847 (31%) | 963/3,400 (28%) | 664/2,176 (31%) | 242/733 (33%) | 162/500 (33%) |
| Over \$100,000 | 2,837/9,998 (28%) | 810/2,342 (35%) | 403/847 (48%) | 572/3,400 (17%) | 650/2,176 (30%) | 218/733 (30%) | 184/500 (37%) |
| Race/Ethnicity |  |  |  |  |  |  |  |
| White, not Hispanic | 6,839/9,998 (68%) | 1,834/2,342 (78%) | 653/847 (77%) | 2,147/3,400 (63%) | 1,301/2,176 (60%) | 553/733 (75%) | 350/500 (70%) |
| Hispanic | 1,572/9,998 (16%) | 265/2,342 (11%) | 98/847 (12%) | 586/3,400 (17%) | 444/2,176 (20%) | 98/733 (13%) | 81/500 (16%) |
| Black | 958/9,998 (9.6%) | 81/2,342 (3.5%) | 24/847 (2.9%) | 507/3,400 (15%) | 270/2,176 (12%) | 42/733 (5.7%) | 34/500 (6.8%) |
| Asian | 183/9,998 (1.8%) | 31/2,342 (1.3%) | 27/847 (3.2%) | 27/3,400 (0.8%) | 73/2,176 (3.3%) | 13/733 (1.7%) | 13/500 (2.6%) |
| Other | 445/9,998<br>(4.5%) | 130/2,342<br>(5.6%) | 44/847 (5.2%) | 132/3,400<br>(3.9%) | 89/2,176 (4.1%) | 27/733 (3.7%) | 22/500 (4.5%) |
| Education |  |  |  |  |  |  |  |
| High School or Less | 4,535/9,998<br>(45%) | 956/2,342 (41%) | 211/847 (25%) | 2,009/3,400<br>(59%) | 953/2,176 (44%) | 282/733 (38%) | 125/500 (25%) |
| Some College | 3,333/9,998<br>(33%) | 838/2,342 (36%) | 335/847 (40%) | 984/3,400 (29%) | 702/2,176 (32%) | 285/733 (39%) | 188/500 (38%) |
| College or More | 2,130/9,998<br>(21%) | 549/2,342 (23%) | 301/847 (36%) | 406/3,400 (12%) | 521/2,176 (24%) | 166/733 (23%) | 186/500 (37%) |
| Employment Status |  |  |  |  |  |  |  |
| Employed | 8,247/9,998<br>(82%) | 1,920/2,342<br>(82%) | 761/847 (90%) | 2,625/3,400<br>(77%) | 1,896/2,176<br>(87%) | 597/733 (81%) | 448/500 (90%) |
| Unemployed | 1,750/9,998<br>(18%) | 422/2,342 (18%) | 86/847 (10%) | 775/3,400 (23%) | 280/2,176 (13%) | 136/733 (19%) | 51/500 (10%) |
| Health Insurance |  |  |  |  |  |  |  |
| Insured | 8,917/9,998<br>(89%) | 2,103/2,342<br>(90%) | 815/847 (96%) | 2,844/3,400<br>(84%) | 2,027/2,176<br>(93%) | 657/733 (90%) | 472/500 (94%) |
| Uninsured | 1,080/9,998<br>(11%) | 239/2,342 (10%) | 32/847 (3.7%) | 556/3,400 (16%) | 150/2,176<br>(6.9%) | 76/733 (10%) | 28/500 (5.6%) |
| Self Reported Health |  |  |  |  |  |  |  |
| Fair/Poor | 806/9,998<br>(8.1%) | 191/2,342<br>(8.1%) | 75/847 (8.9%) | 273/3,400<br>(8.0%) | 172/2,176<br>(7.9%) | 51/733 (7.0%) | 44/500 (8.7%) |
| Good | 2,503/9,998<br>(25%) | 597/2,342 (26%) | 237/847 (28%) | 811/3,400 (24%) | 497/2,176 (23%) | 209/733 (28%) | 152/500 (30%) |
| Very good | 3,628/9,998<br>(36%) | 881/2,342 (38%) | 315/847 (37%) | 1,131/3,400<br>(33%) | 827/2,176 (38%) | 287/733 (39%) | 188/500 (38%) |
| Excellent | 3,061/9,998<br>(31%) | 673/2,342 (29%) | 220/847 (26%) | 1,185/3,400<br>(35%) | 681/2,176 (31%) | 186/733 (25%) | 116/500 (23%) |
| Religious Status |  |  |  |  |  |  |  |
| Religious | 7,870/9,998<br>(79%) | 1,812/2,342<br>(77%) | 670/847 (79%) | 2,612/3,400<br>(77%) | 1,776/2,176<br>(82%) | 581/733 (79%) | 419/500 (84%) |
| Atheist/Agnostic | 2,127/9,998<br>(21%) | 530/2,342 (23%) | 177/847 (21%) | 787/3,400 (23%) | 400/2,176 (18%) | 152/733 (21%) | 81/500 (16%) |
| Have child age 5 to 11 | 6,214/9,998<br>(62%) | 1,347/2,342<br>(58%) | 549/847 (65%) | 2,084/3,400<br>(61%) | 1,451/2,176<br>(67%) | 436/733 (59%) | 348/500 (70%) |
| Have child age 12 to 15 | 4,725/9,998<br>(47%) | 1,257/2,342<br>(54%) | 405/847 (48%) | 1,586/3,400<br>(47%) | 858/2,176 (39%) | 396/733 (54%) | 223/500 (45%) |
| Have child age 16 to 17 | 2,792/9,998<br>(28%) | 784/2,342 (33%) | 228/847 (27%) | 962/3,400 (28%) | 481/2,176 (22%) | 237/733 (32%) | 100/500 (20%) |
| Party |  |  |  |  |  |  |  |
| Republican | 4,767/9,998<br>(48%) | 1,236/2,342<br>(53%) | 487/847 (58%) | 1,459/3,400<br>(43%) | 986/2,176 (45%) | 339/733 (46%) | 260/500 (52%) |
| Democrat | 1,231/9,998<br>(12%) | 102/2,342<br>(4.4%) | 48/847 (5.6%) | 502/3,400 (15%) | 444/2,176 (20%) | 69/733 (9.4%) | 67/500 (13%) |
| Independent | 4,000/9,998<br>(40%) | 1,004/2,342<br>(43%) | 312/847 (37%) | 1,439/3,400<br>(42%) | 747/2,176 (34%) | 325/733 (44%) | 173/500 (35%) |
| When will the<br>Pandemic end? |  |  |  |  |  |  |  |
| Already Over | 4,492/9,998<br>(45%) | 1,513/2,342<br>(65%) | 506/847 (60%) | 1,344/3,400<br>(40%) | 653/2,176 (30%) | 320/733 (44%) | 157/500 (31%) |
| Less<br>than three<br>months | 560/9,998<br>(5.6%) | 66/2,342 (2.8%) | 38/847 (4.5%) | 201/3,400<br>(5.9%) | 174/2,176<br>(8.0%) | 45/733 (6.1%) | 35/500 (7.1%) |
| Less<br>than one year | 1,213/9,998<br>(12%) | 162/2,342<br>(6.9%) | 64/847 (7.6%) | 495/3,400 (15%) | 323/2,176 (15%) | 108/733 (15%) | 61/500 (12%) |
| More<br>than one year | 3,732/9,998<br>(37%) | 601/2,342 (26%) | 239/847 (28%) | 1,359/3,400<br>(40%) | 1,027/2,176<br>(47%) | 261/733 (36%) | 246/500 (49%) |
| Flu vaccine since<br>6/21 | 2,111/9,998<br>(21%) | 189/2,342<br>(8.1%) | 314/847 (37%) | 345/3,400 (10%) | 971/2,176 (45%) | 68/733 (9.3%) | 224/500 (45%) |
| Parent<br>Vaccination<br>Status |  |  |  |  |  |  |  |
| Unvaccinated | 6,475/9,998<br>(65%) | 2,342/2,342<br>(100%) | 0/847 (0%) | 3,400/3,400<br>(100%) | 0/2,176 (0%) | 733/733 (100%) | 0/500 (0%) |
| Partially<br>Vaccinated | 818/9,998<br>(8.2%) | 0/2,342 (0%) | 190/847 (22%) | 0/3,400 (0%) | 529/2,176 (24%) | 0/733 (0%) | 99/500 (20%) |
| Fully Vaccinated | 2,223/9,998<br>(22%) | 0/2,342 (0%) | 569/847 (67%) | 0/3,400 (0%) | 1,339/2,176<br>(62%) | 0/733 (0%) | 315/500 (63%) |
| Fully Vaccinated<br>and Boosted | 482/9,998<br>(4.8%) | 0/2,342 (0%) | 88/847 (10%) | 0/3,400 (0%) | 309/2,176 (14%) | 0/733 (0%) | 85/500 (17%) |
| Parent COVID-19<br>Vaccination Type |  |  |  |  |  |  |  |
| Johnson |  |  | 175/847 (21%) |  | 317/2,176 (15%) |  | 68/500 (14%) |
| mRNA |  |  | 672/847 (79%) |  | 1,859/2,176<br>(85%) |  | 431/500 (86%) |
| Local Adverse<br>Effect |  |  | 679/847 (80%) |  | 1,539/2,176<br>(71%) |  | 417/500 (83%) |
| Systemic Adverse<br>Effect |  |  | 622/847 (73%) |  | 1,245/2,176<br>(57%) |  | 403/500 (81%) |
| Rash |  |  | 120/847 (14%) |  | 182/2,176<br>(8.4%) |  | 60/500 (12%) |
| Nausea/Vomiting |  |  | 130/847 (15%) |  | 184/2,176<br>(8.5%) |  | 77/500 (15%) |
| Muscle Ache |  |  | 439/847 (52%) |  | 709/2,176 (33%) |  | 299/500 (60%) |
| Pain in Arm |  |  | 662/847 (78%) |  | 1,444/2,176 (66%) |  | 411/500 (82%) |
| Swelling in Arm |  |  | 239/847 (28%) |  | 368/2,176 (17%) |  | 100/500 (20%) |
| Fever |  |  | 266/847 (31%) |  | 497/2,176 (23%) |  | 183/500 (37%) |
| Chills |  |  | 291/847 (34%) |  | 521/2,176 (24%) |  | 214/500 (43%) |
| Tiredness |  |  | 497/847 (59%) |  | 891/2,176 (41%) |  | 325/500 (65%) |
| Headache |  |  | 393/847 (46%) |  | 692/2,176 (32%) |  | 267/500 (53%) |
| Something Else |  |  | 105/847 (12%) |  | 197/2,176 (9.1%) |  | 64/500 (13%) |

\*All Parents includes parents who are unvaccinated, partially vaccinated, fully vaccinated, and fully vaccinated and boosted
\*\*Vaccinated parents includes parents who are partially vaccinated, fully vaccinated, or fully vaccinated and boosted

In addition, 73% of parents in Cluster 1 are unvaccinated, while 61% and 59% of parents in Clusters 2 and 3 are unvaccinated, respectively. However, we find that the three types of vaccinated parents (partially vaccinated, fully vaccinated, fully vaccinated and boosted) are relatively evenly distributed across the three clusters.

## Discussion

In this study, we aimed to identify the determinants of parental COVID-19 vaccine hesitancy and the main motivating beliefs for reluctance to vaccinate children among hesitant parents. We found parental vaccination status to be a large predictor of parent preference for child vaccination. In addition, we find heterogeneity among parents both in correlates of willingness and reasons for hesitancy. For instance, several correlates differed by parental vaccination status. Among the unvaccinated, younger age and lower education were associated with willingness, and among the vaccinated an inverse association was observed. Reasons for hesitancy to vaccinate children also differed by parental vaccination status with unvaccinated parents less likely to trust government and scientists. Hesitant parents can be divided into three categories based on their reasons for hesitancy. To our knowledge, this is the first study to use a cluster analysis on parental vaccine hesitancy. Additionally, previous research has suggested that the survey is representative of the larger population [25].

Our findings on the correlates of willingness to vaccinate children against COVID-19 are generally consistent with previous studies. Previous research has found vaccinated parents much more likely than unvaccinated parents to be willing to vaccinate their children [8, 12-22]. Prior studies have also found likelihood of child vaccination higher among parents of older children, older parents, higher educated parents, Democrat-affiliated parents, insured parents, higher income parents, Hispanic and Asian parents, parents with routine influenza vaccine behavior, and male gender [12, 13, 15, 21 22, 28, 29]. Our non-stratified results are generally consistent with prior work. Unlike past research, we find lower income parents have higher odds for willingness on all three multivariate models. We additionally find an inverse relationship between willingness and higher age and education among unvaccinated parents.

The WHO’s 3 Cs (confidence, complacency, and convenience) model of vaccine hesitancy is an oft used mechanism to model barriers to vaccine uptake [30]. Prior research on parental COVID-19 vaccine hesitancy have found the top reasons are often confidence related, including vaccine safety and effectiveness, side effects, and lack of trust in government [12-15]. We find consistent results with past studies in the prevalence of concerns over vaccine safety, side effects, and lack of trust in government. However, we find that for a majority of parents (Cluster 2), side effects rarely factor into the decision to vaccinate children, suggesting complacency or convenience considerations as the main factors driving hesitancy among this group. Simply looking at the top concerns then, will ignore important distinctions. Moreover, parents in Cluster 1 and Cluster 3 both have severe confidence concerns, with large portions in both groups concerned about side effects and the speed at which the vaccine was developed. Yet a large majority of Cluster 1 parents exhibit a lack of trust in government or scientists while a small minority of Cluster 3 parents exhibit a lack of trust in both. Treating parents with side effect concerns as a homogenous group may ignore important differences and may fail to sufficiently ease their concerns. Policymakers, public health organizations, and physicians must better tailor their messages to these disparate groups in order to increase vaccination rates among children.

Several limitations must be considered in interpreting the study results. First, bias is expected from any observational study conducted using a convenience sample; those willing to participate in our study may not be representative of the public as a whole and our findings may not be generalizable to the broader US population. The use of survey weights and the anonymity of the survey may help lessen this bias. Second, we did not have direct measures of certain variables of interest such as child COVID-19 vaccination status and instead must rely on self-report. However, previous research on this survey’s estimates have shown it to track closely with CDC estimates for overall vaccination rates as well as alternative estimates for pandemic behavior such as mask wearing across the US [24, 31]. Third, results may be limited by the dynamic nature of the COVID-19 pandemic with changes in public policy, public health recommendations, and case prevalence during the study period. For instance, parents today may express different preferences towards vaccinations as newer options become more available such as Novavax and new variants circulate.

Willingness to vaccinate children is associated with many different parental characteristics with parent vaccination status being a major determinant. Further work is needed to tailor child vaccination efforts to subgroups of parents who are more hesitant. It is important to acknowledge that predictors of parental vaccine hesitancy differ among vaccinated and unvaccinated parents and that examining hesitant parents’ concerns in aggregate will obscure many important distinctions.

## Data Availability

Data used for all analyses, tables, and figures available here: https://github.com/sehgal-neil/parental-vaccine-hesitancy

## Acknowledgments

The authors thank Kara Sewalk and the SurveyMonkey research team (Laura Wronski, Tim Gravelle & Jon Cohen) for their assistance. Dr. Astley acknowledges funding from NIH/NIDDK: K23 DK120899. Ben Rader, Autumn Gertz & Dr. Brownstein acknowledge funding from the Centers for Disease Control and Prevention. The funding organizations had no role in the design and conduct of the study; collection, management, analysis, and interpretation of the data; preparation, review, or approval of the manuscript; and decision to submit the manuscript for publication. Mr. Sehgal had full access to all the data in the study and takes responsibility for the integrity of the data and the accuracy of the data analysis.

## Tables and Figures

**Appendix Table 1.**
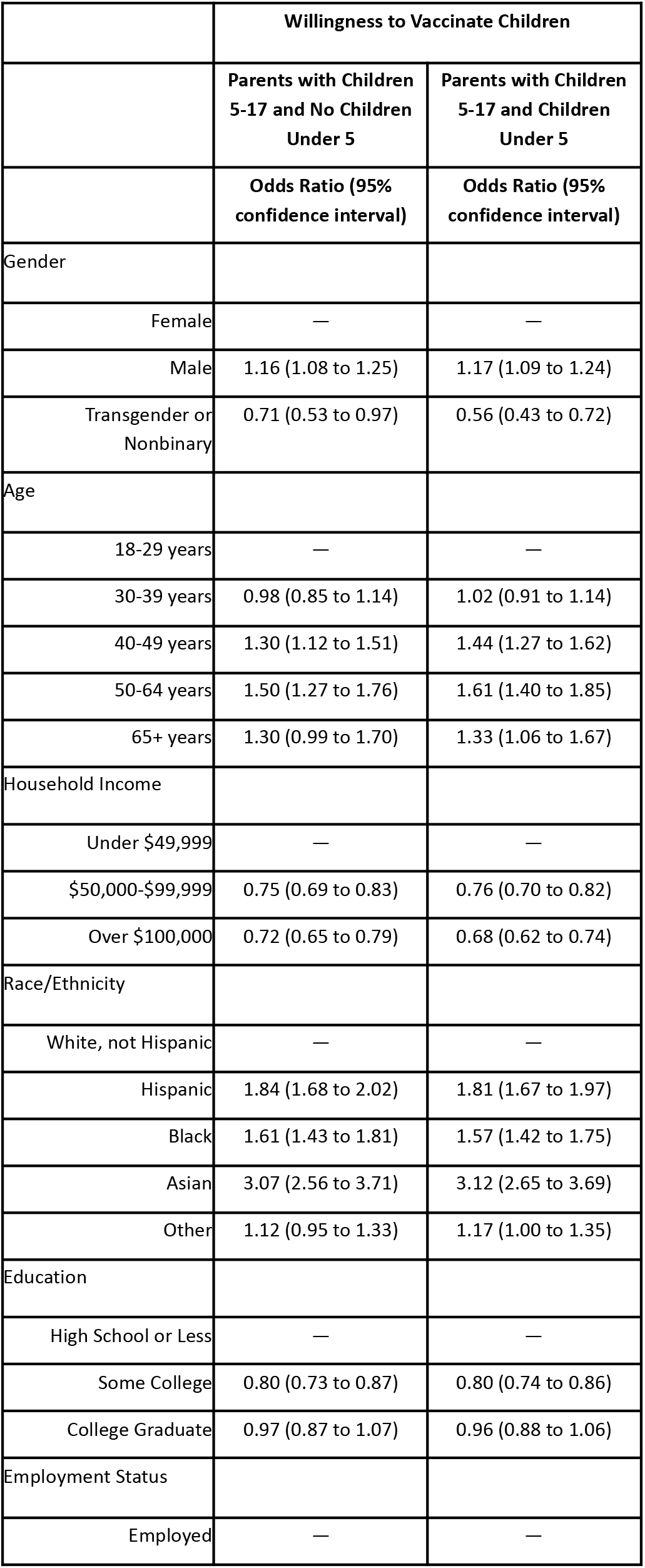
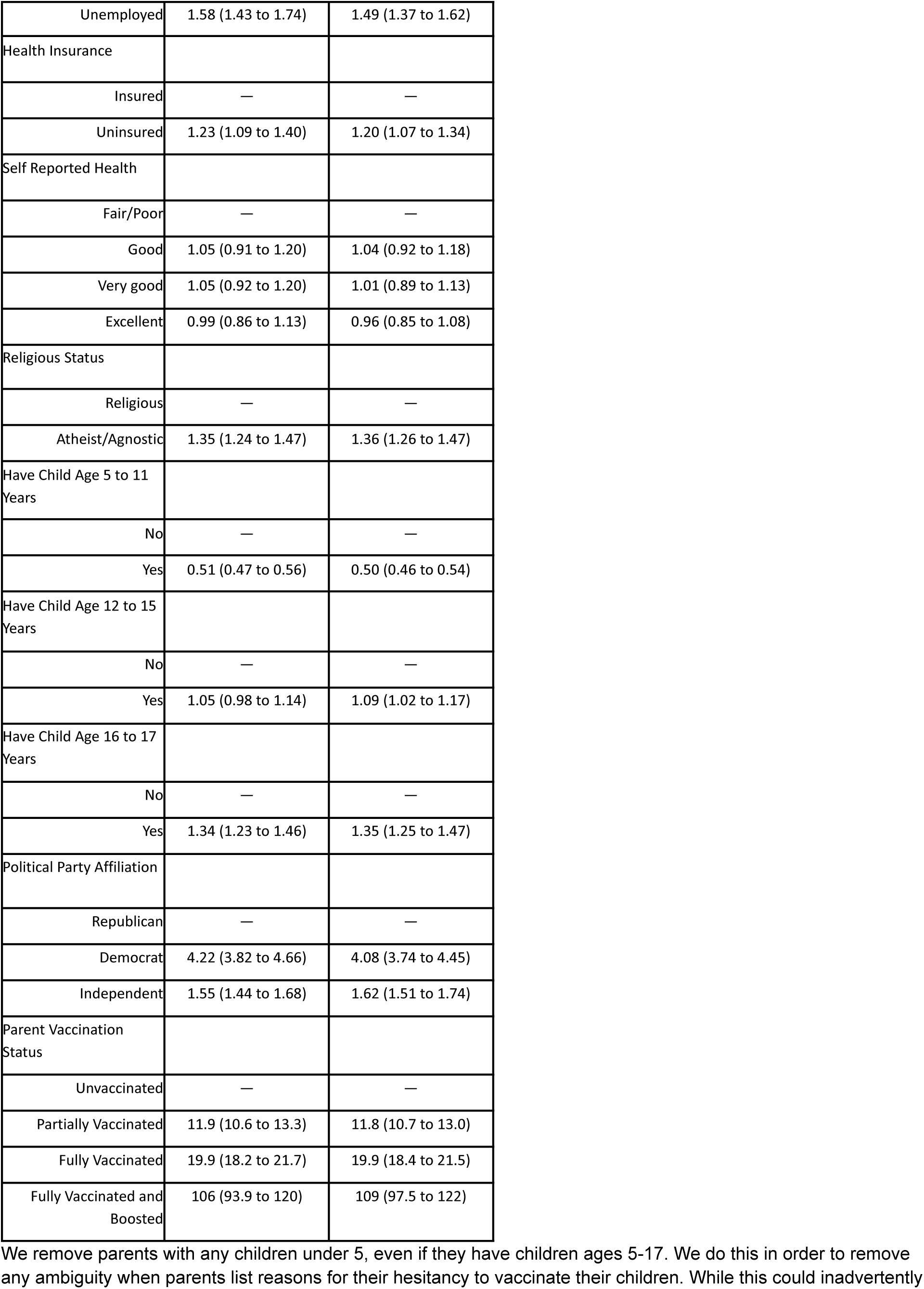
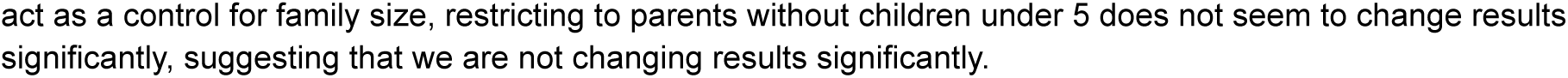
Multivariate Results for Parents with Children Under 5 and No Children Under 5

**Appendix Table 2.** Reasons for Hesitancy by Cluster

|  | *All Hesitant Parents | Cluster 1 |  | Cluster 2 |  | Cluster 3 |  |
| --- | --- | --- | --- | --- | --- | --- | --- |
|  | Overall, N = 9,998 | Unvaccinated, N = 2,342 | Vaccinated**, N = 847 | Unvaccinated, N = 3,400 | Vaccinated**, N = 2,176 | Unvaccinated, N = 733 | Vaccinated**, N = 500 |
| **Reason** |  |  |  |  |  |  |  |
| Child received flu vaccine and is protected | 55/9,998 (0.5%) | 33/2,342 (1.4%) | 0/847 (0%) | 20/3,400 (0.6%) | 0/2,176 (0%) | 2/733 (0.3%) | 0/500 (0%) |
| Vaccine too new | 4,350/9,998 (44%) | 1,912/2,342 (82%) | 645/847 (76%) | 454/3,400 (13%) | 334/2,176 (15%) | 605/733 (83%) | 400/500 (80%) |
| Child too young | 2,295/9,998 (23%) | 808/2,342 (35%) | 373/847 (44%) | 357/3,400 (10%) | 386/2,176 (18%) | 202/733 (28%) | 169/500 (34%) |
| Let child decide when older | 2,040/9,998 (20%) | 790/2,342 (34%) | 292/847 (35%) | 377/3,400 (11%) | 303/2,176 (14%) | 170/733 (23%) | 109/500 (22%) |
| Side Effects | 4,673/9,998 (47%) | 1,946/2,342 (83%) | 686/847 (81%) | 502/3,400 (15%) | 450/2,176 (21%) | 651/733 (89%) | 438/500 (88%) |
| COVID-19 threat exaggerated | 2,403/9,998 (24%) | 1,434/2,342 (61%) | 476/847 (56%) | 266/3,400 (7.8%) | 135/2,176 (6.2%) | 52/733 (7.1%) | 40/500 (8.0%) |
| Lack trust in government | 3,963/9,998 (40%) | 2,175/2,342 (93%) | 746/847 (88%) | 680/3,400 (20%) | 228/2,176 (10%) | 101/733 (14%) | 34/500 (6.7%) |
| Lack trust in scientists | 3,385/9,998 (34%) | 1,924/2,342 (82%) | 690/847 (81%) | 446/3,400 (13%) | 201/2,176 (9.2%) | 82/733 (11%) | 42/500 (8.4%) |
| Vaccine development too political | 3,546/9,998 (35%) | 2,016/2,342 (86%) | 738/847 (87%) | 365/3,400 (11%) | 245/2,176 (11%) | 109/733 (15%) | 73/500 (15%) |
| Child already had COVID-19 | 2,525/9,998 (25%) | 880/2,342 (38%) | 281/847 (33%) | 549/3,400 (16%) | 406/2,176 (19%) | 264/733 (36%) | 146/500 (29%) |
| Child never gets any vaccine | 719/9,998 (7.2%) | 290/2,342 (12%) | 26/847 (3.1%) | 349/3,400 (10%) | 40/2,176 (1.8%) | 13/733 (1.8%) | 1/500 (0.2%) |
| Vaccine not recommended for child's health history | 507/9,998 (5.1%) | 185/2,342 (7.9%) | 73/847 (8.6%) | 92/3,400 (2.7%) | 80/2,176 (3.7%) | 46/733 (6.3%) | 30/500 (6.1%) |
| Worried child will get COVID-19 from vaccine | 622/9,998 (6.2%) | 357/2,342 (15%) | 55/847 (6.4%) | 103/3,400 (3.0%) | 44/2,176 (2.0%) | 48/733 (6.5%) | 16/500 (3.3%) |
| Child is not at risk | 1,662/9,998 (17%) | 747/2,342 (32%) | 255/847 (30%) | 260/3,400 (7.6%) | 221/2,176 (10%) | 101/733 (14%) | 79/500 (16%) |
| Risk from vaccine greater than risk from COVID-19 | 3,601/9,998 (36%) | 1,743/2,342 (74%) | 613/847 (72%) | 333/3,400 (9.8%) | 165/2,176 (7.6%) | 443/733 (60%) | 303/500 (61%) |
| Child is afraid of needles | 479/9,998 (4.8%) | 176/2,342 (7.5%) | 38/847 (4.5%) | 97/3,400 (2.9%) | 89/2,176 (4.1%) | 40/733 (5.5%) | 37/500 (7.4%) |
| Vaccine is contrary to religious beliefs | 1,101/9,998 (11%) | 587/2,342 (25%) | 56/847 (6.6%) | 312/3,400 (9.2%) | 33/2,176 (1.5%) | 106/733 (14%) | 7/500 (1.4%) |
| Prefer to wait for herd immunity for protection | 1,360/9,998 (14%) | 645/2,342 (28%) | 251/847 (30%) | 171/3,400 (5.0%) | 137/2,176 (6.3%) | 92/733 (13%) | 63/500 (13%) |
| There are other people who should get it first | 223/9,998 (2.2%) | 76/2,342 (3.2%) | 25/847 (2.9%) | 30/3,400 (0.9%) | 59/2,176 (2.7%) | 17/733 (2.3%) | 17/500 (3.5%) |
| Other | 939/9,998 (9.4%) | 217/2,342 (9.3%) | 70/847 (8.2%) | 356/3,400 (10%) | 229/2,176 (11%) | 39/733 (5.3%) | 29/500 (5.7%) |
| Median Number of Reasons Given for Hesitancy | 3.0 (1.0 – 6.0) | 8.0 (6.0 – 10.0) | 7.0 (6.0 – 9.0) | 1.0 (1.0 – 2.0) | 1.0 (1.0 – 2.0) | 4.0 (3.0 – 5.0) | 4.0 (3.0 – 5.0) |

\*All Parents includes parents who are unvaccinated, partially vaccinated, fully vaccinated, and fully vaccinated and boosted
\*\*Vaccinated parents includes parents who are partially vaccinated, fully vaccinated, or fully vaccinated and boosted

**Appendix Table 3.**
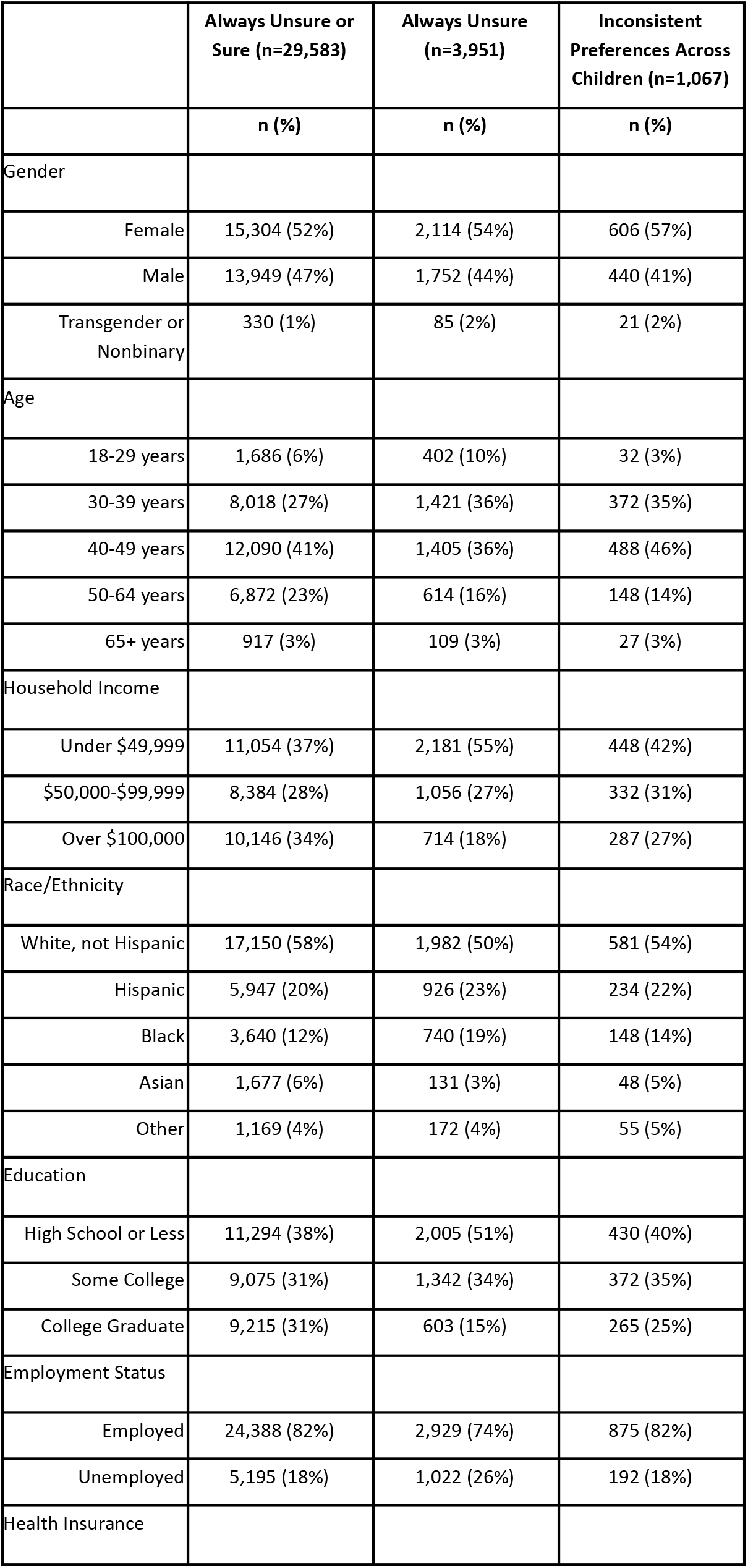
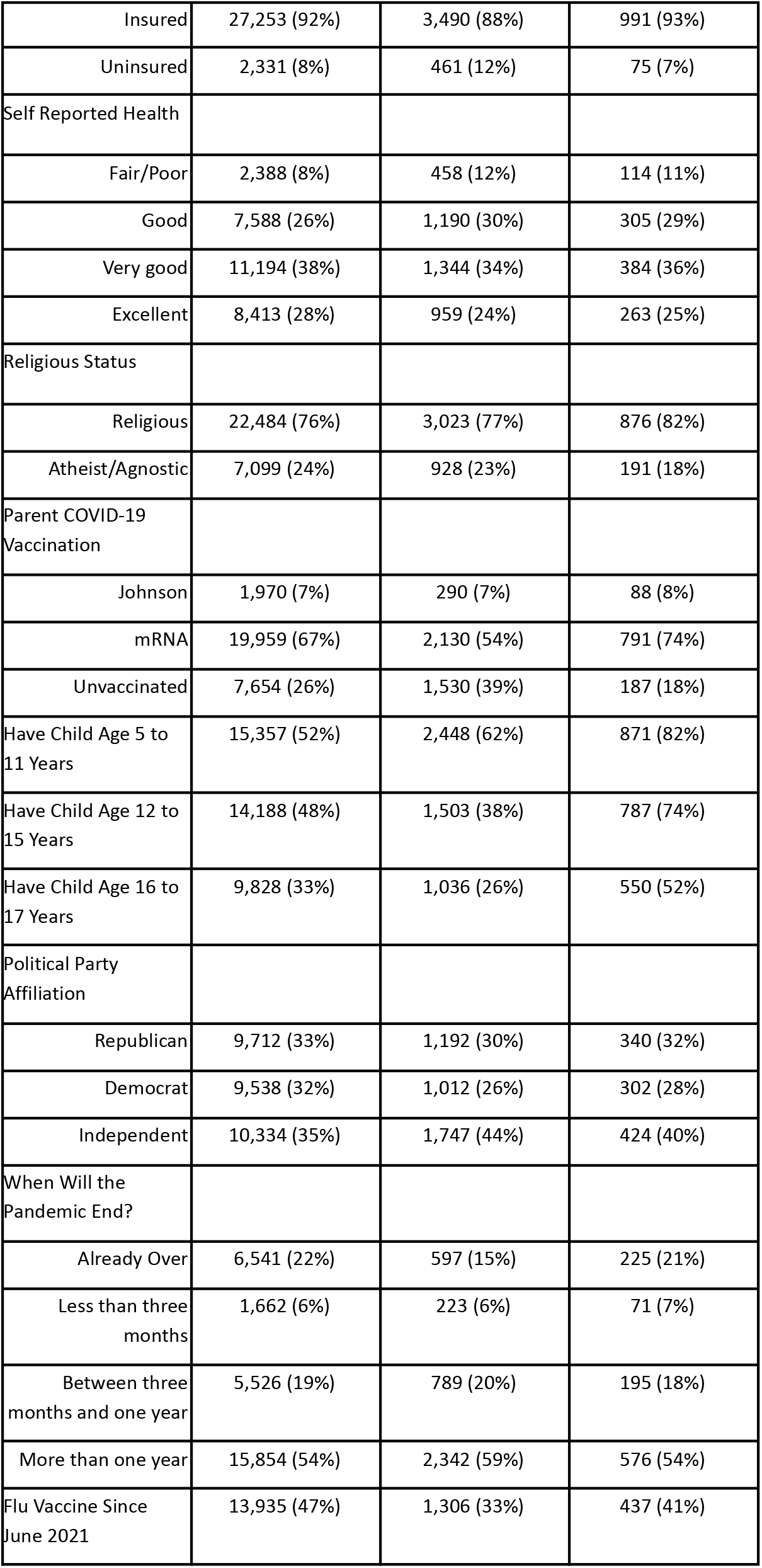
Characteristics of Parents with Uncertain or Inconsistent Preferences

**Appendix Table 4.**
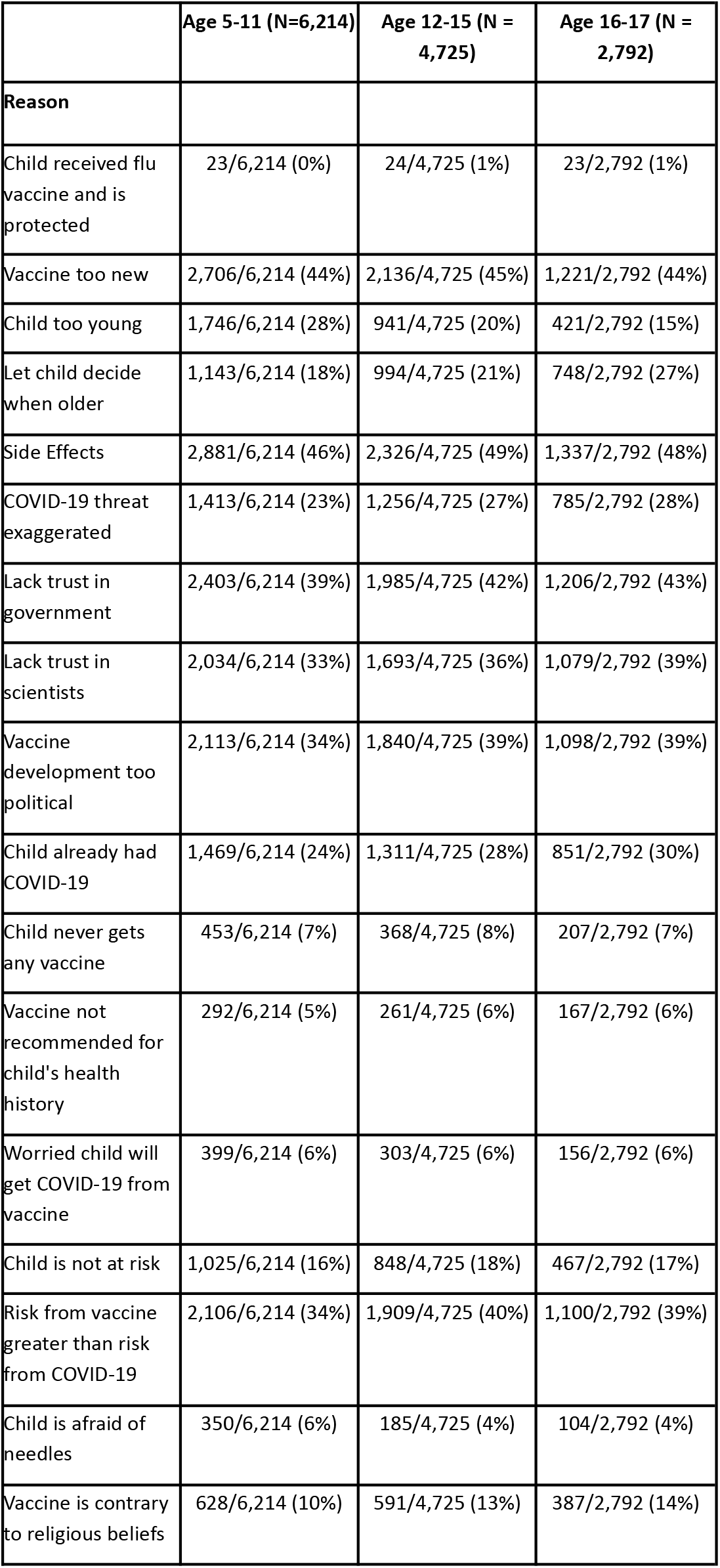
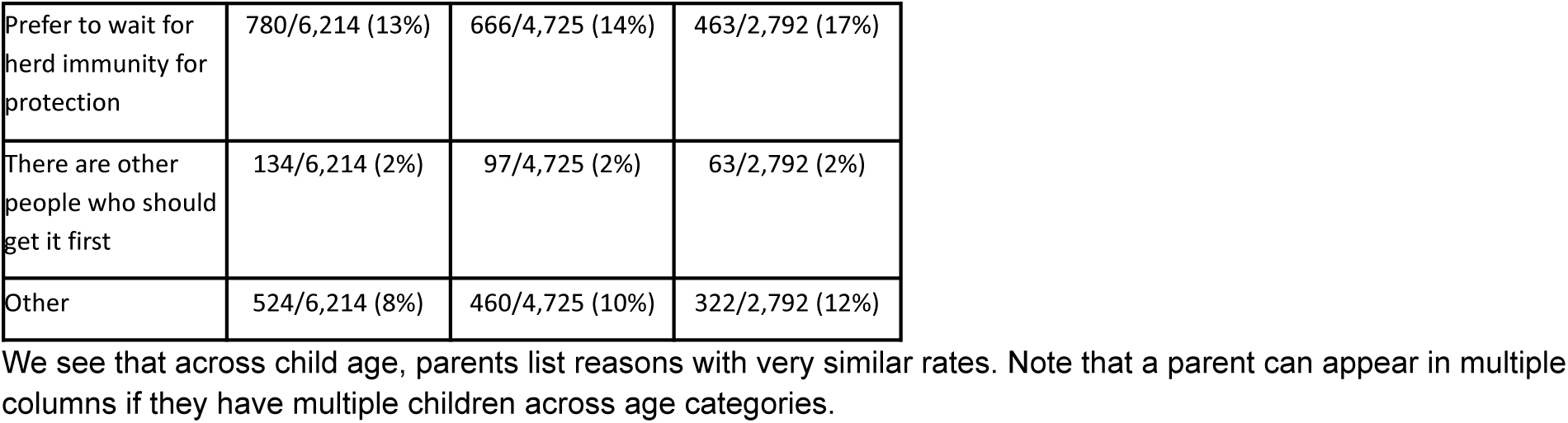
Reasons for Hesitancy by Child Age

|  | Age 5-11 (N=6,214) | Age 12-15 (N = 4,725) | Age 16-17 (N = 2,792) |
| --- | --- | --- | --- |
| **Reason** |  |  |  |
| Child received flu vaccine and is protected | 23/6,214 (0%) | 24/4,725 (1%) | 23/2,792 (1%) |
| Vaccine too new | 2,706/6,214 (44%) | 2,136/4,725 (45%) | 1,221/2,792 (44%) |
| Child too young | 1,746/6,214 (28%) | 941/4,725 (20%) | 421/2,792 (15%) |
| Let child decide when older | 1,143/6,214 (18%) | 994/4,725 (21%) | 748/2,792 (27%) |
| Side Effects | 2,881/6,214 (46%) | 2,326/4,725 (49%) | 1,337/2,792 (48%) |
| COVID-19 threat exaggerated | 1,413/6,214 (23%) | 1,256/4,725 (27%) | 785/2,792 (28%) |
| Lack trust in government | 2,403/6,214 (39%) | 1,985/4,725 (42%) | 1,206/2,792 (43%) |
| Lack trust in scientists | 2,034/6,214 (33%) | 1,693/4,725 (36%) | 1,079/2,792 (39%) |
| Vaccine development too political | 2,113/6,214 (34%) | 1,840/4,725 (39%) | 1,098/2,792 (39%) |
| Child already had COVID-19 | 1,469/6,214 (24%) | 1,311/4,725 (28%) | 851/2,792 (30%) |
| Child never gets any vaccine | 453/6,214 (7%) | 368/4,725 (8%) | 207/2,792 (7%) |
| Vaccine not recommended for child's health history | 292/6,214 (5%) | 261/4,725 (6%) | 167/2,792 (6%) |
| Worried child will get COVID-19 from vaccine | 399/6,214 (6%) | 303/4,725 (6%) | 156/2,792 (6%) |
| Child is not at risk | 1,025/6,214 (16%) | 848/4,725 (18%) | 467/2,792 (17%) |
| Risk from vaccine greater than risk from COVID-19 | 2,106/6,214 (34%) | 1,909/4,725 (40%) | 1,100/2,792 (39%) |
| Child is afraid of needles | 350/6,214 (6%) | 185/4,725 (4%) | 104/2,792 (4%) |
| Vaccine is contrary to religious beliefs | 628/6,214 (10%) | 591/4,725 (13%) | 387/2,792 (14%) |
| Prefer to wait for herd immunity for protection | 780/6,214 (13%) | 666/4,725 (14%) | 463/2,792 (17%) |
| There are other people who should get it first | 134/6,214 (2%) | 97/4,725 (2%) | 63/2,792 (2%) |
| Other | 524/6,214 (8%) | 460/4,725 (10%) | 322/2,792 (12%) |

We see that across child age, parents list reasons with very similar rates. Note that a parent can appear in multiple columns if they have multiple children across age categories.

